# Relative role of border restrictions, case finding and contact tracing in controlling SARS-CoV-2 in the presence of undetected transmission

**DOI:** 10.1101/2021.05.05.21256675

**Authors:** Rachael Pung, Hannah E. Clapham, Vernon J. Lee, Adam J Kucharski, on behalf of the CMMID COVID-19 working group

## Abstract

**Background:** Several countries have controlled the spread of COVID-19 through varying combinations of border restrictions, case finding, contact tracing and careful calibration on the resumption of domestic activities. However, evaluating the effectiveness of these measures based on observed cases alone is challenging as it does not reflect the transmission dynamics of missed infections.

**Methods:** Combining data on notified local COVID-19 cases with known and unknown sources of infections (i.e. linked and unlinked cases) in Singapore in 2020 with a transmission model, we reconstructed the incidence of missed infections and estimated the relative effectiveness of different types of outbreak control. We also examined implications for estimation of key real-time metrics — the reproduction number and ratio of unlinked to linked cases, using observed data only as compared to accounting for missed infections.

**Findings:** Prior to the partial lockdown in Singapore, initiated in April 2020, we estimated 89% (95%CI 75–99%) of the infections caused by notified cases were contact traced, but only 12.5% (95%CI 2–69%) of the infections caused by missed infectors were identified. We estimated that the reproduction number was 1.23 (95%CI 0.98–1.54) after accounting for missed infections but was 0.90 (95%CI 0.79-1.1) based on notified cases alone. At the height of the outbreak, the ratio of missed to notified infections was 34.1 (95%CI 26.0–46.6) but the ratio of unlinked to linked infections was 0.81 (95%CI 0.59–1.36). Our results suggest that when case finding and contact tracing identifies at least 50% and 20% of the infections caused by missed and notified cases respectively, the reproduction number could be reduced by more than 14%, rising to 20% when contact tracing is 80% effective.

**Interpretation:** Depending on the relative effectiveness of border restrictions, case finding and contact tracing, unobserved outbreak dynamics can vary greatly. Commonly used metrics to evaluate outbreak control — typically based on notified data — could therefore misrepresent the true underlying outbreak.

**Funding:** Ministry of Health, Singapore.

**Research in context:** *Evidence before this study:* We searched PubMed, BioRxiv and MedRxiv for articles published in English up to Mar 20, 2021 using the terms: (2019-nCoV OR “novel coronavirus” OR COVID-19 OR SARS-CoV-2) AND (border OR travel OR restrict* OR import*) AND (“case finding” OR surveillance OR test*) AND (contact trac*) AND (model*). The majority of modelling studies evaluated the effectiveness of various combinations of interventions in the absence of outbreak data. For studies that reconstructed the initial spread of COVID-19 with outbreak data, they further simulated counterfactual scenarios in the presence or absence of these interventions to quantify the impact to the outbreak trajectory. None of the studies disentangled the effects of case finding, contact tracing, introduction of imported cases and the reproduction number, in order to reproduce an observed SARS-CoV-2 outbreak trajectory.

*Added value of this study:* Notified COVID-19 cases with unknown and known sources of infection are identified through case finding and contact tracing respectively. Their respective daily incidence and the growth rate over time may differ. By capitalising on these differences in the outbreak data and the use of a mathematical model, we could identify the key drivers behind the growth and decline of both notified and missed COVID-19 infections in different time periods — e.g. domestic transmission vs external introductions, relative role of case finding and contact tracing in domestic transmission. Estimating the incidence of missed cases also allows us to evaluate the usefulness of common surveillance metrics that rely on observed cases.

*Implications of all the available evidence:* Comprehensive outbreak investigation data integrated with mathematical modelling helps to quantify the strengths and weaknesses of each outbreak control intervention during different stages of the pandemic. This would allow countries to better allocate limited resources to strengthen outbreak control. Furthermore, the data and modelling approach allows us to estimate the extent of missed infections in the absence of population wide seroprevalence surveys. This allows us to compare the growth dynamics of notified and missed infections as reliance on the observed data alone may create the illusion of a controlled outbreak.

## Background

The COVID-19 pandemic has resulted in substantial disruption to international travel and trade due to widespread border restrictions that have been enacted by countries as a key strategy to reduce the importation and spread of COVID-19.^1^ In addition to border controls, which do not totally prevent the importation of cases,^2^ case finding and contact tracing have formed a central part of the response in many countries. Case finding has helped in early identification and isolation of new infections that are not associated with other known cases through testing of suspected cases and surveillance in target groups.^3,4^ Meanwhile, contact tracing identifies potential transmission routes and new infections among contacts of a known case.^3,4^ Local COVID-19 cases with known and unknown sources of infection can therefore be categorised as ‘linked’ or ‘unlinked’ respectively.^3,5^

The occurrence of unlinked COVID-19 cases implies that the pandemic is partly unobserved. This could be attributed to the importation and transmission from asymptomatic or mildly symptomatic infections who do not require medical attention^6^ and underreporting of symptomatic cases^7^. Furthermore, failure to trace secondary cases arising from a notified case would also create gaps in the observed transmission chains. The respective case counts and ratio of unlinked to linked cases are often used as a metric for the effectiveness of outbreak control, and are closely monitored in many countries to assess the potential for resuming social and economic activities and the lifting of border restrictions.^8–10^ However, methods to establish the relative role of border restrictions, case finding, contact tracing to the trajectory of linked and unlinked cases, remain elusive.

In addition, the pandemic trajectory is often measured by the effective reproduction number which relies on notified cases or cases extrapolated from observed deaths.^11–13^ With effective case isolation and contact tracing, the time spent in the community while infectious for a notified case is truncated, shortening the observed serial interval (a proxy for generation interval).^14^ Consequently, the observed chains of transmission are short-lived and, on average, less than one secondary case is generated.^15^ As such, it is currently unclear whether metrics based on observed features such as notified cases and their linkage give an accurate picture on the underlying outbreak dynamics.

As countries progressively reopen, it is important for policy makers to understand the parameters that contributes to the spread of cases and the growth dynamics in undetected cases. Combining the daily incidence of imported, and local linked and unlinked COVID-19 cases in Singapore with a mathematical model, we aim to characterise the effectiveness of detection and control measures over the course of the pandemic and estimate the incidence of missed cases, even when longitudinal serology surveys are absent. Furthermore, we compared the growth patterns in missed and notified cases and investigated the implications of making inferences on the pandemic trajectory based on observed data alone.

## Methods

### Data

Cases of COVID-19 (confirmed with a respiratory sample positive for SARS-CoV-2 on PCR) notified to the Ministry of Health, Singapore from Jan 23 to Dec 31, 2020 in Singapore were used. Extensive epidemiological investigations are conducted for each case to establish their exposure history prior to notification.^16–19^ A local linked case was a case with at least one known source of infection while a local unlinked case was a case with an unknown source of infection. Case finding measures (i.e. active surveillance) identify local unlinked cases through testing of persons with acute respiratory infection or pneumonia, routine testing of targeted groups at high-risk of acquiring or transmitting disease, and ad-hoc testing of sub-populations of interest.^3^

Imported cases are confirmed COVID-19 cases with travel history to a country with ongoing COVID-19 outbreak in the preceding 14 days. They were stratified into two main categories, those quarantined in dedicated facilities upon arrival, and those undergoing home-based quarantine or who were not quarantined at all, prior to detection.^20^ The former were tested at the start and end of their quarantine period and were assumed to be incapable of introducing infections into the community while the latter, to whom the community were potentially exposed to, could generate local infections.

All confirmed cases were conveyed to secured isolation facilities and discharged after 21 days from date of confirmation if assessed to be clinically well, or with sequential negative tests. Cases occurring in persons residing in a foreign-worker dormitory and notified from Apr 7 to Oct 31, 2020 were omitted from analysis as these dormitories were placed under lockdown for an extended period of time. As workers were subjected to restricted movements, the opportunity to interact with the community during this period was minimal and hence they were assumed to be incapable of driving community-level transmission. Furthermore, about 0.2% of the confirmed cases occurred in persons providing care to confirmed cases and as these secondary infections were not community-acquired infections, they were omitted from analysis.

### Transmission Model

We simulated disease transmission through a branching process to compute the expected incidence over time. The model was fitted using a Poisson likelihood to the daily incidence of linked and unlinked local cases to reconstruct the incidence of missed infections (see Supplementary Information for details).

Infections were introduced into the population by either notified imported cases who potentially had contact with community individuals (referred to as notified imported cases with community contact in the remaining text) or through missed imported infections. We modelled the number of missed imported infections relative to the number of notified imported cases with community contact using a factor *ρ*. Both types of imported cases could generate community infections from the time of arrival to isolation or end of their infectiousness respectively. Community infections were identified through varying effectiveness of contract tracing of notified cases (i.e. probability of detecting linked cases, *ε*_link_) or case finding (i.e. probability of detecting unlinked cases, *ε*_unlink_). The potential reproduction number of an infected individual, *R*, was defined as the average number of secondary cases generated by a single infectious individual over the entire infectious period in the absence of quarantine/isolation. This is analogous to the reproduction number of a missed infected individual, *R*_missed_. For a notified case, the amount of time spent in the community while infectious is generally shorter as compared to a missed infection, either as they sought medical attention and were isolated, or when a secondary case was identified through contact tracing and quarantined before being tested positive. The reproduction number of a notified case, *R*_notified_, was defined as the average number of secondary cases generated by a single infectious individual till the time of quarantine/isolation. Notified cases were assumed to be incapable of generating offspring infections once isolated. Overall, the effective reproduction number, *R*_eff_, is an aggregate measure of the reproduction number of both missed and notified cases.

We assumed the generation time was gamma distributed with mean 7.5 days (SD 3.4).^21^ The distribution was left-truncated for all imported cases to exclude the infectious period while overseas and right-truncated for all notified cases due to early case isolation.

Due to varying effectiveness of detection and control measures, and changing social behaviour over the course of the pandemic, we defined five different time periods with a different value estimated for the four unknown parameters (*ρ, ε*_link_, *ε*_unlink_, *R*) for each period – when transmission was mainly driven by (i) travellers from China, and (ii) travellers from other countries with ongoing outbreak; the implementation of (iii) partial lockdown within Singapore; (iv) resumption of local activities, and (v) increased reopening of national borders. Results for *ρ* were converted to the average daily number of missed imported cases for each time period to provide an estimate of the magnitude of missed imported infections and we report the posterior median with 95% credible intervals (CI) for all model outputs.

### Independent model validation

We validated the model outcomes against two independent population level surveys involving (i) 41,852 preschool staff who underwent a PCR test between May 15 and 29, 2020 followed by serological testing for confirmed cases and (ii) 1,578 participants of a cross-sectional seroprevalence survey conducted from Oct 7 to 31, 2020 with participants randomly selected from the general population.^22^ Incorporating PCR detection and seroconversion probabilities,^23,24^ we estimated PCR and serology positive cases from the model to compare against the incidence rate in preschool staff, and seroconverted cases from the model to compare against the seroprevalence rate in the general population (Supplementary Information). Observed data were presented as the mean and the 95% confidence intervals for binomial proportions were computed using Wilson’s method.^25^ We performed a z-test to evaluate the difference between the observed and modelled rates and p values <0.05 were considered statistically significant.

### Modelling varying effectiveness of detecting linked and unlinked cases and impact to epidemic growth dynamics

We further explored the epidemic growth dynamics in the presence of missed and notified SARS-CoV-2 infections via the use of a next-generation matrix mathematical framework (Supplementary Information). In general, a notified case spends a reduced amount of time spent in the community while infectious as compared to a missed infection. This creates a heterogeneity in the reproduction number of a missed and notified case. As such, we studied the impact of the *R*_eff_ across a range of values for *ε*_link_, *ε*_unlink_ and *R*. We assumed a Weibull distributed time of infection to isolation with mean 9.2 days (SD 4.4) for notified cases (derived from observed data in symptomatic cases notified from Mar 1 to Apr 6, 2020, Supplementary Figure 1 and Table 1).

The ratio of missed to notified cases established during exponential growth was also modelled to characterise the extent of case ascertainment and compared against the ratio of unlinked to linked infections. Lastly, we analysed how different ratios of missed to notified imported cases with community contact influences the time taken to achieve exponential growth and implications of inferring outbreak trajectory from transient growth patterns.

All data and code required to reproduce the analysis is available online.^26^

## Results

In Singapore, initial border and outbreak control measures from Jan 18 to Feb 29, 2020 aimed to reduce the spread of SARS-CoV-2 by infected persons arriving from China. These measures were progressively expanded from Mar 1, 2020 in response to a surge of imported cases returning from countries with ongoing outbreaks (figure 2a). Despite the decline in notified imported cases from Mar 16 to Apr 1, 2020, community transmissions were rising (figure 2b and c). By the time a nationwide partial lockdown was implemented on Apr 7, 2020, 810 community infections had been notified. However, our model estimated the majority of the infections had gone undetected, with 2,940 (95% CI 414–26,500) missed (figure 2d). During the partial lockdown period and Phase 1 of reopening (Apr 7–Jun 18, 2020), the estimated daily number of missed cases decreased but remained above 100 and cumulatively, 20,000 infections (95% CI 15,200–34,400) were missed (figure 2d). As border restrictions were gradually lifted and economic and social activities resumed, the number of imported cases with community contact were kept low and there was no evidence of sustained community transmission (figure 2a–c). Overall, we estimated that 26,700 cases (95% CI 19,400–64,000) were missed in 2020 (figure 2d).

**Figure 1.**
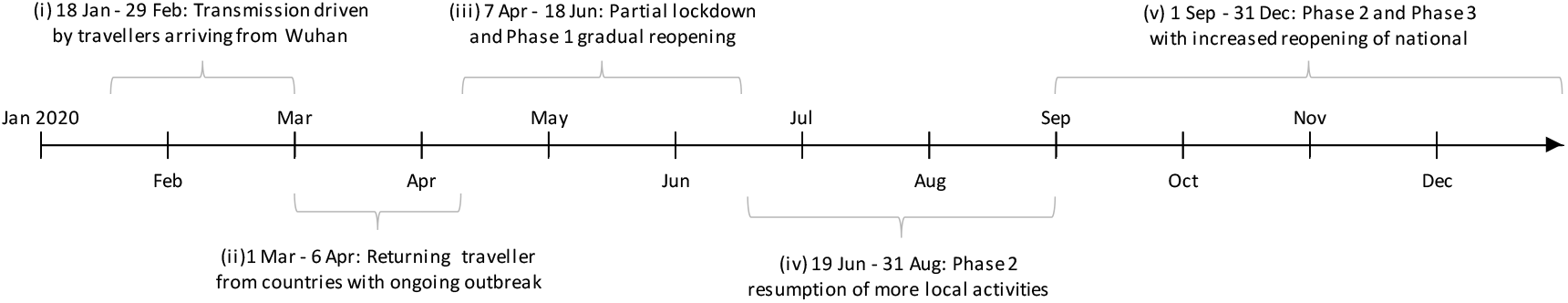
Time periods considered in transmission model

**Figure 2.**
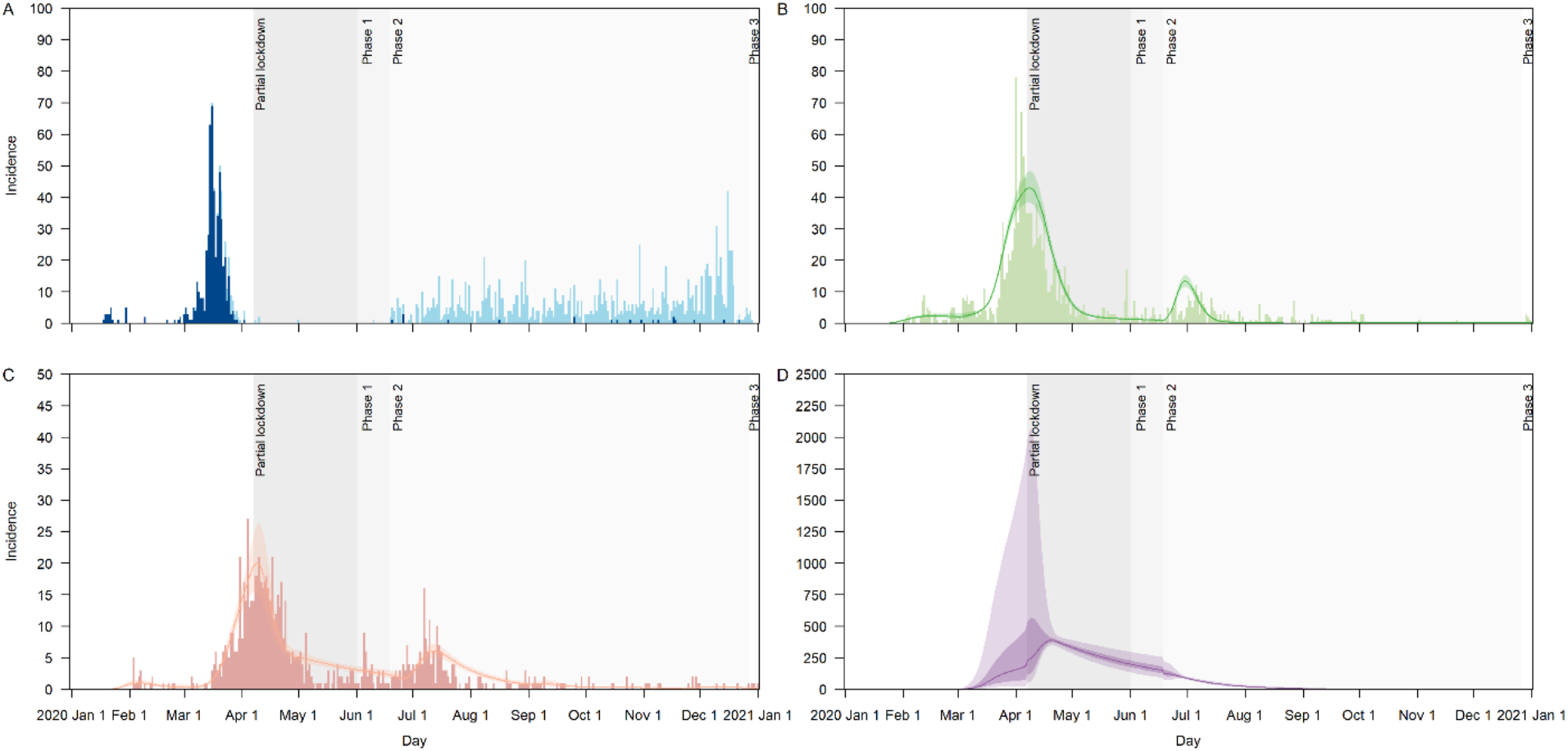
Daily incidence of COVID-19 cases in Singapore, (A) notified imported cases with community contact (dark blue bars) and notified imported cases who were quarantined upon arrival (light blue bars), (B) notified local linked cases (green bars) and the posterior median (green line) and 95% CI (green shaded area), (C) notified local unlinked cases (pink bars) and the posterior median (pink line) and 95% CI (pink shaded area), (D) modelled posterior median local missed infections (line), 50% CI (dark purple shaded area) and 95% CI (light purple shaded area).

We estimated that *R* was 1.17 (95% CI 0.97–1.35) at the start of the pandemic. From Mar 1 to Apr 6, 2020, this increased to 1.38 (95% CI 1.21–1.67) prior to the partial lockdown (figure 3a). However, *R*_notified_, was lower at 0.90 (95% CI 0.79–1.1) due to the reduced amount of time spent in the community while infectious compared to a missed infection (supplementary figure 5).. After accounting for missed infections, the *R*_eff_ was 1.23 (95% CI 0.98–1.54) (supplementary figure 5). While the country’s contact tracing system was able to detect 89% of the secondary cases (*ε*_link_, 95% CI 75–99%) arising from a notified case, only 12.5% of the infections (*ε*_unlink_, 95%CI 2-69%) caused by a missed infected individual were identified through case finding — a sharp decline from 78% (*ε*_unlink_, 95% CI 37–99%) as estimated from Jan 18 to Feb 29 (figure 3b and c). We observed a peak of 70 imported cases per day being isolated and we estimated that 23 imported cases (95% CI 3–175) were missed daily (figure 3d).

**Figure 3.**
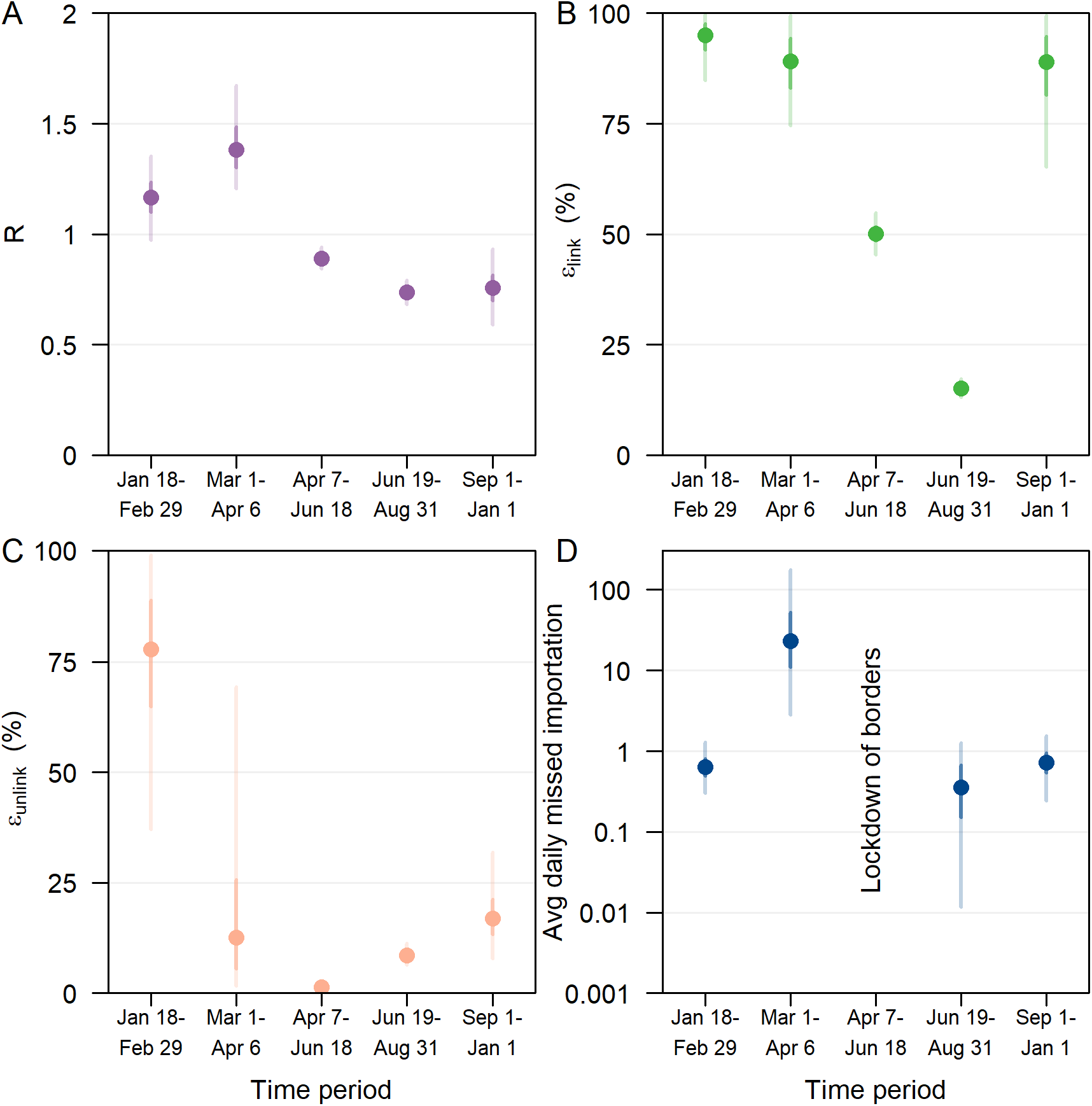
Posterior median (dot), 50% CI (dark vertical lines) and 95% CI (light vertical lines) of model parameters. (A) potential reproduction number, *R*, of a COVID-19 case, (B) effectiveness of detecting a linked case, *ε*_link_, (C) effectiveness of detecting an unlinked case, *ε*_unlink_, (D) average daily number of missed imported cases in log scale.

During the partial lockdown, although we estimated *R* to be below 1, which signalled a controlled outbreak, the effectiveness of detecting linked and unlinked cases was low at 50% (95%CI 45–55%) and 1% (95%CI 1–2%) respectively (figure 3b and c). As such, it took about two months to reach a daily observed incidence of less than 10 (figure 2b and c). As social and economic activities were progressively resumed from Jun 19, 2020 onwards, the effectiveness of detecting linked and unlinked cases remained low at 15% (95% CI 13–17%) and 8% (95% CI 6–11%) respectively. However, with strict quarantine of incoming travellers and continued enforcement of outbreak control measures, the average daily number of missed imported infections was low at 0.35 (95% CI 0.01–1.27) (figure 3c and d) and *R* was approximately 0.74 (95% CI 0.68–0.79) (figure 3a and d). By Sep 1, 2020 the contact tracing system’s effectiveness in identifying secondary cases from a notified case had recovered to 89% (95%CI 65–99%) (figure 3b).

### Independent model validation

While the transmission model was able to reproduce the observed temporal trends, we sought to further validate the model outputs against independent population level surveys. Firstly, of the 41,852 preschool staff tested between May 15 and 29, 2020, 13 staff were PCR and serology positive for SARS-CoV-2 infection, which translates to an observed incidence rate of 311 infections per million population (95%CI 181–531) compared to our model estimate of 355 per million population (95%CI 302–421). Secondly, in a cross-sectional seroprevalence survey from Oct 7 to 31, 2020, SARS-CoV-2 antibodies were detected in 7 out of 1,578 participants.^20^ The observed seroprevalence was 0.44% (95%CI 0.22–0.91%) compared to our model estimate of 0.43% (95%CI 0.31–1.03%). The difference between the observed and modelled rates in both surveys were statistically insignificant (p=0.63 and p=0.93).

### Varying effectiveness of detecting linked and unlinked cases and implications for epidemic growth

When the effectiveness of detecting linked cases is low (*ε*_link_=20%), an effectiveness of detecting unlinked cases of at least 50% results in *R*_eff_ approximately 14.1% lower than *R* (figure 4). As the testing capacity and effectiveness of case finding increases, with a further increase in the ability to ring fence secondary infections arising from notified cases, *ε*_link_, of up to 80%, this could further reduce the *R*_eff_ to at least 23.3% lower than *R* (figure 4). As a country’s case finding and contact tracing system strengthens and the time from infection to isolation is reduced, the reduction in the *R*_eff_ relative to *R* is expected to increase (supplementary figure 6 and 7).

**Figure 4.**
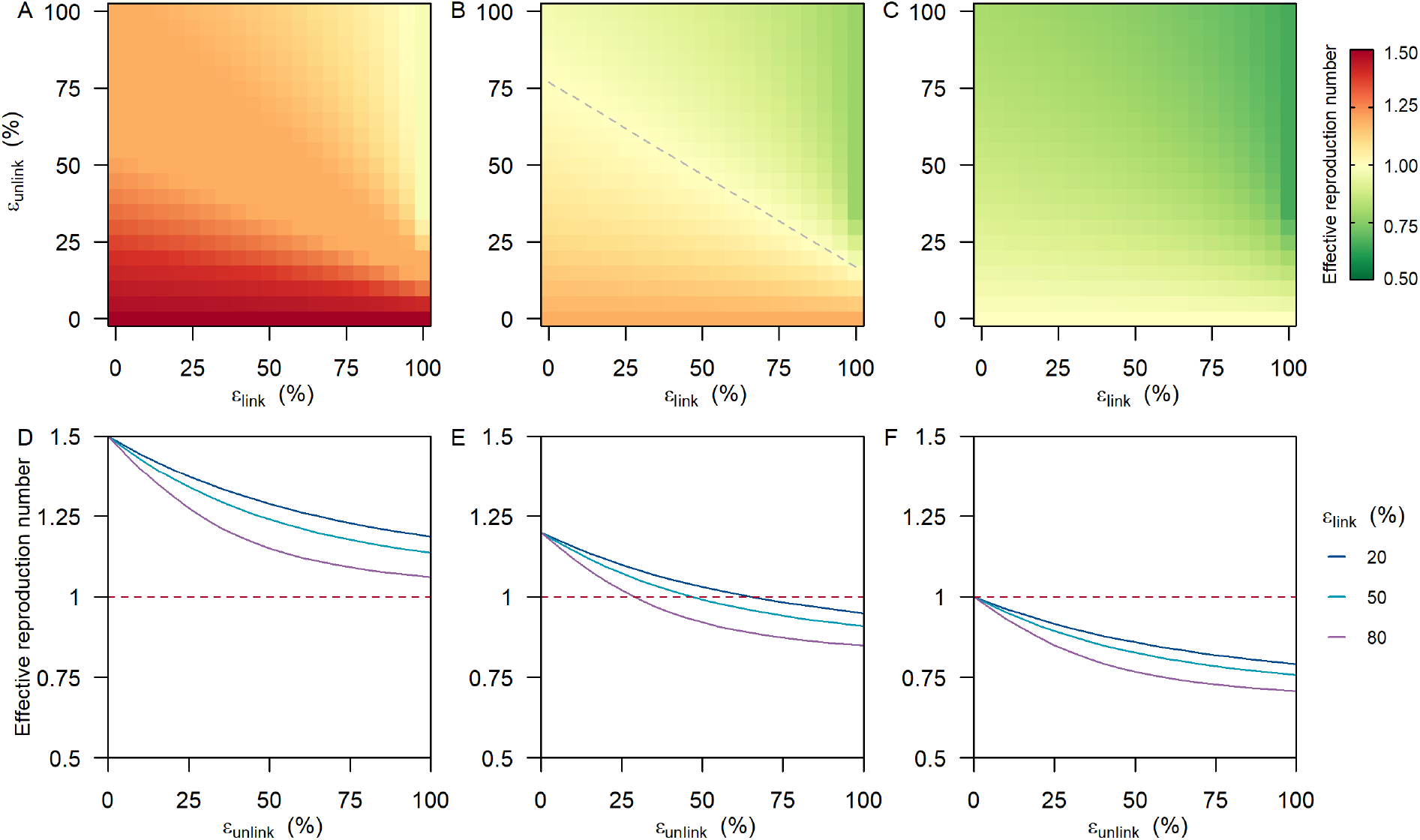
Effect of detecting linked and unlinked cases on the effective reproduction number, *R*_eff_. For a potential reproduction number, *R* of 1.5 (A, D), 1.2 (B, E) and 1.0 (C, F) and a fixed distribution of time from infection to isolation (Weibull distributed with mean 9.2 days (SD 4.4)), increasing levels of *ε*_link_ and *ε*_unlink_ creates a heterogenous pool of missed and notified infections with the latter being subjected to early case isolation and hence lowers the *R*_eff_. (B) Grey dashed line represents *R*_eff_ of 1.

To characterise the extent of case ascertainment, we estimate the ratio of missed to notified cases. When *ε*_link_ is more than 80%, this ratio remains below 1 if *ε*_unlink_ is greater than 30%. In other words, aggressive ring fencing of contacts whenever a confirmed case is detected, helps to ensure that the underlying outbreak is mostly observed (figure 5a). For the same values of *ε*_link_ and *ε*_unlink_, the ratio of missed to notified cases can be vastly different from the ratio of unlinked to linked cases — typically used to characterise the extent of outbreak control. When *ε*_unlink_ is low, the epidemic becomes increasingly obscure and the ratio of missed to notified cases grows very large but the ratio of the unlinked to linked cases tends to a fixed value dependent on *ε*_link_ (figure 5a and b).

**Figure 5.**
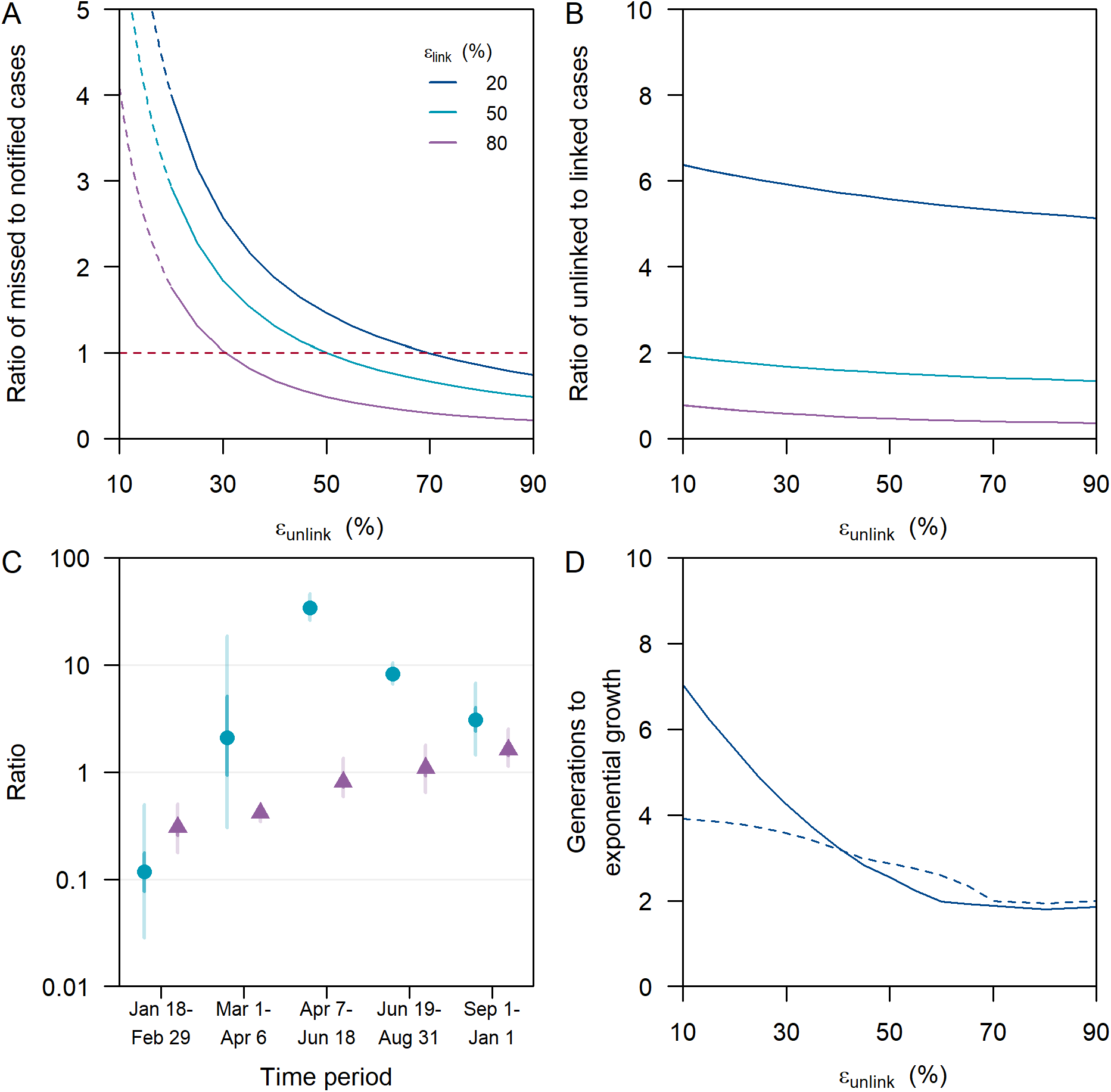
Comparison of unobserved outbreak dynamics with common surveillance metrics. (A) Ratio of missed to notified cases for varying *ε*_link_ and *ε*_unlink_. Dashed lines implies that the ratio extends to infinity as *ε*_unlink_ tends to zero; (B) Ratio of unlinked to linked cases for varying *ε*_link_ and *ε*_unlink_; (C) Ratio of missed to notified cases (turquoise dots) and ratio of unlinked to linked cases (purple triangle) in different time periods of the pandemic. Posterior median (dot/triangle), 50% CI (dark vertical lines) and 95% CI (light vertical lines). (D) Generations to exponential growth for an outbreak with 5 missed imported cases and 95 notified imported cases with community contact, *R* of 1.5, *ε*_link_ of 80%, for varying *ε*_unlink_.

To put these results into the Singapore context, as the effectiveness of detecting unlinked cases declined in March during the surge of imported cases and further declined during the partial lockdown, the ratio of missed to notified cases increased to 34.1 (95%CI 26.0–46.6) and was many times higher than the ratio of unlinked to linked cases of 0.81 (95%CI 0.59–1.36) (figure 5c). As such, metrics derived from observed data alone do not always accurately reflect the underlying outbreak.

Under certain conditions, the time to achieve exponential growth in notified and missed infections could differ. When *ε*_unlink_ is low and when the initial ratio of missed to notified imported cases with community contact is low (5 missed:95 notified), the number of generations required to achieve exponential growth in notified cases is greater than that in missed cases (figure 5d). This time to exponential growth is reduced when either the *ε*_unlink_ is high or when the initial ratio of missed to notified imported cases with community contact increases (figure 5d and supplementary figure 8).

## Discussion

Using the growth patterns in the daily incidence of local linked and unlinked cases, and imported cases with community contact, our model was capable of disentangling the effects of case finding and contact tracing (*ε*_unlink_ and *ε*_link_) from other outbreak interventions that affect the potential reproduction number of a case (*R*), at a time before vaccination roll out. In spite of a strong capability to contact trace, without a tight control on the number of imported cases coupled with low ability to detect new cases and *R*_eff_ exceeding 1, community transmission was sustained, as witnessed in Singapore’s daily incidence of COVID-19 cases from Mar 1 to Apr 6, 2020. This surge in community cases affected the ability of the contact tracing system to ring fence notified cases in the following months (figure 3b) but the partial lockdown with strong enforcement of non-pharmaceutical interventions such as mask wearing, physical distancing, and movement restrictions helped to reverse the pandemic trajectory (figure 2 and 3).

As countries progressively resume economic and social activities in partially vaccinated populations, the potential reproduction number of an infectious individual engaged in these activities may exceed unity. Case finding and contact tracing help in the early identification and isolation of (secondary) cases thereby minimising the duration of infectious period spent in the community. Even if contact tracing capacity is low, if more than half of the infections arising from a previously undetected infection present to the healthcare system for early testing and isolation, the *R* of a case could be reduced by more than 14% (figure 4). Increasing the effectiveness of case finding (*ε*_unlink_) implies casting a wider surveillance net but ultimately this measure depends on compliance with testing regimes. We estimated a sharp drop in *ε*_unlink_ during the partial lockdown period (figure 3c) and this behaviour was corroborated by behavioural surveys documenting diminished health-seeking behaviour.^27^ Factors driving avoidance of testing warrants further studies as, no matter how many infections can be detected from contact tracing, the healthcare system relies on the testing and identification of cases in the first instance.

As contact tracing devices are progressively rolled out to speed up contact tracing and coupled with effective quarantine and testing of close contacts, for a given *ε*_unlink_ of 50% and *ε*_link_ of 80%, *R* is lowered by more than 20% (figure 4). Even if a healthcare system is only able to detect 30–50% of the infections arising from previously undetected infections (*ε*_unlink_), a high *ε*_link_ ensures aggressive ring fencing of secondary infections arising from an unlinked case and keeps the outbreak in check (figure 5a).

Current methods to estimate the effective reproduction number, *R*_eff_, of SARS-CoV-2 rely on the notified case incidence or deaths, some with appropriate adjustments to capture the right-censoring of data.^11–13,15^ However, in the presence of asymptomatic infections^6^ and underreporting of symptomatic cases^7^, these methods neglect the growth dynamics of a non-negligible number of missed infections which can result in misleading inferences. From the model, we estimated that *R* was about 1.38 from Mar 1 to Apr 6, 2020 (figure 3a). With an effective contact tracing system that quarantines close contacts and detects close to 90% of the secondary cases from notified cases, this results in reduced time from infection to isolation and the reproduction number of a notified case was 0.90, similar to previous modelling estimates.^15^ Collectively, the *R*_eff_ was approximately 1.2, which exceeds 1 and signalled sustained transmissions (supplementary figure 5).

By reconstructing the daily incidence of missed infections, we could infer the time-varying level of case under-ascertainment. We estimated that nearly 90% of the missed infections occurred between Mar 1 and Jun 18, 2020 and the ratio of missed to notified infections was more than 30 (figure 5c). This overall level of case under-ascertainment was comparable to the estimated under-ascertainment in symptomatic cases alone in other countries during similar phases of the pandemic and the under-ascertainment levels in many countries are expected to be higher when accounting for asymptomatic infections.^7^ Furthermore, we showed that using the ratio of unlinked to linked cases — calculated from observed data alone, does not adequately characterise the extent of outbreak control. The ratio of unlinked to linked cases showed little variation over the course of the pandemic as compared to the ratio of missed to notified infections and does not provide warning of a runaway outbreak.

With the reopening of borders, missed and notified imported cases with community contact could initiate the first generation of local infections and if there is a low ratio of missed to notified imported case of 1:19 coupled with a low ability to detect unlinked cases (*ε*_unlink_ = 10%), it takes 3 more generations for the notified cases to achieve exponential growth as compared to the missed cases (figure 5d). The deviation from exponential growth in notified cases at the early stages of the outbreak creates an illusion that outbreak can be controlled based on observed data, while the number of undetected infections continues to escalate. One effective but resource intensive method of minimising the number of missed imported infections is to implement strict quarantine and testing of incoming travellers, especially those arriving from countries with high COVID-19 incidence. This reduces the overall number of new infections introduced into the population and is especially important for countries that have managed to stabilise their epidemic numbers after the initial wave(s) of infections.

There are some limitations to our study. Firstly, the model assumes that each of the four parameters remains constant in a specified time period. As such, we are unable to provide a time-varying measure to characterise the impact of different outbreak detection and control measures that were progressively rolled out in the population at a granular level. Instead, time periods were chosen based on prior knowledge of major policies that would affect at least one of the four model parameters. Secondly, those imported cases subjected to home quarantine and no quarantine were assumed to have the same potential of making contact with members of the community as household transmission might occur, while in reality the amount of community contact would be different as those under home quarantine could potentially only contact household members. In the absence of data, we made a conservative assumption that the imported cases under home quarantine were capable of generating local infections. Further model calibration would require data on the outcome of the various quarantine measures. Finally, missed infections could arise from asymptomatic or mildly symptomatic infections, or underreporting of symptomatic cases. We assumed that *R* is the same among these cases. More information is needed to determine the temporal variation in the types of cases to account for lowered transmission potential in asymptomatic or mildly symptomatic cases. At the same time, a key strength to our analysis is that our model was able to reproduce independent observations in two separate population level surveys and this lends support to our assumption of a homogeneous *R* among all missed infections.

The SARS-CoV-2 pandemic has generated new forms of data collection and many new ways to reconstruct outbreak dynamics and evaluate the extent of missed infections arising from high asymptomatic rates and underreporting of cases. The daily incidence of linked and unlinked cases could help countries evaluate their performance in case finding, contact tracing and the effectiveness of their border restrictions. Missed and notified infections bring about a heterogeneity in the reproduction number and the mixture of these factors can create an illusion of a controlled outbreak. As countries progressively reopen borders or plan for pandemics in the future, it is important to have an integrated surveillance and modelling analysis system to overcome the challenges of undetected transmissions.

## Data Availability

All data and code required to reproduce the analysis is available online at https://github.com/rachaelpung/covid_missed_infections

