## supplementary for "Relative role of border restrictions, case finding and contact tracing in controlling SARS-CoV-2 in the presence of undetected transmission"

[+] List of members given at end of Appendix

### Modelling the incidence of COVID-19 infections

In the model, all notified infections will be isolated and infections were initiated by the following types of imported infectors:

- Notified imported cases, $n_{import}$: individuals infected overseas, arrived on time $t_a$, had exposure to the community and were isolated by time $t_i$. The time of infection, $t_\tau$, and consequently, the duration from infection to arrival, $a$ (i.e. $t_a - t_\tau$), is unknown. As such, we derive the marginal probability distribution of the duration of infection to arrival, $\alpha$, based on the observed distribution of time from arrival to symptoms onset in notified, symptomatic imported cases and the incubation period distribution for SAR-CoV-2 infections.[1] For each notified imported case, we then derive the distribution of infection times using this margin probability distribution and $t_a$.

- Missed imported infections, $m_{import}$: individuals infected overseas, arrived at time $t_a$, had exposure to the community and were never isolated. We applied a smoothing spline to the number of notified imported case with arrival time $t_a$ and had contact with the community and modelled the number of missed imported case arriving on $t_a$ as $\boldsymbol{\rho}$ times of the smoothing spline of notified imported case with arrival time $t_a$.

  Similar to the notified imported case, we derived the distribution of infection times for missed imported cases using the margin probability distribution of the duration of infection to arrival and $t_a$.

At any given time $t$, the number of secondary infections (i.e. offspring infections) generated in the community $I_{comm}$ at time $t$ by notified imported cases prior to isolation and missed imported cases is:

$$I_{comm}(t) = \int_0^\tau \int_0^\infty \left[n_{import}(t-\tau, a) + m_{import}(t-\tau, a)\right] \beta(\tau) d\tau da \quad (1)$$

where $n_{import}(t-\tau,a)$ are individuals infected overseas at $t-\tau$, arrived in Singapore at $t-\tau+a$, has a known isolation time but yet to be isolated at time $t$. $m_{import}(t-\tau,a)$ are individuals infected overseas at $t-\tau$, arrived in Singapore at $t-\tau+a$ and never isolated. $\beta(\tau)$ is the mean rate of generating offspring infections by an infector at time $\tau$ since infection. $\beta(\tau)$ is a function of the generation interval, $\omega(\tau)$, which is assumed to be gamma distributed with mean 7.5 days (SD 3.4)[2] and, $R$, the potential reproduction number, defined as the average number of secondary cases generated by a single infectious individual over the course of the entire infectious period in the absence of quarantine/isolation.

The offspring infections generated by the imported cases at time $t$, $I_{comm}(t)$, serves as the next generation of infectors and can be further stratified into notified and missed infectors, $n_{comm}(t)$ and $m_{comm}(t)$ respectively, derived as follows:

$$\begin{aligned} n_{comm}(t) &= \left[\int_0^{\tau}\int_0^{\infty} n_{import}(t-\tau,a)\,\beta(\tau)d\tau da\right]\varepsilon_{link} \\ &+ \left[\int_0^{\tau}\int_0^{\infty} m_{import}(t-\tau,a)\,\beta(\tau)d\tau da\right]\varepsilon_{unlink} \\ &= n_{comm,link}(t) + n_{comm,unlink}(t) \end{aligned} \tag{2}$$

$$\begin{aligned} m_{comm}(t) &= \left[\int_0^{\tau}\int_0^{\infty} n_{import}(t-\tau,a)\,\beta(\tau)d\tau da\right](1-\varepsilon_{link}) \\ &+ \left[\int_0^{\tau}\int_0^{\infty} m_{import}(t-\tau,a)\,\beta(\tau)d\tau da\right](1-\varepsilon_{unlink}) \end{aligned} \tag{3}$$

Equation 2 describes how offspring infections were identified with varying efficacy through contract tracing or through case finding. The former serves to detect offspring infections arising from a notified parent infector (i.e. probability of detecting linked cases, $\varepsilon_{link}$) while the latter detects offspring infections arising from a missed parent infector (i.e. probability of detecting unlinked cases, $\varepsilon_{unlink}$).

Subsequent generations of infections infected at time $t$ are describe by the renewal equation:

$$I_{comm}(t) = \int_0^{\infty}[n_{comm}(t-\tau)\Lambda(\tau) + m_{comm}(t-\tau)]\,\beta(\tau)d\tau \tag{4}$$

where $n_{comm}(t-\tau)$ are individuals infected in the community at $t-\tau$ and yet to be isolated and $m_{comm}(t-\tau)$ are individuals infected in the community at $t-\tau$ but never isolated. We derive the marginal probability distribution of the duration of infection to isolation for community cases, $\gamma$, by convolving the observed distribution of time from symptoms onset in notified, symptomatic community cases to isolation, and the incubation period distribution for SAR-CoV-2 infections.[1] We then derive the probability that a notified case remains at large in the community $\tau$ time since infection, $\Lambda(\tau)$, by taking $1-\int_0^{\tau}\gamma(i)di$.

The new generations of detected offspring infections $n_{comm,link}(t)$ and $n_{comm,unlink}(t)$ arising from $n_{comm}(t-\tau)$ and $m_{comm}(t-\tau)$ follows the same principle as equation (2).

**Likelihood of detecting offspring infections**

$n_{comm,link}(t)$ and $n_{comm,unlink}(t)$ denotes linked and unlinked cases infected at time $t$. The time from infection to isolation follows the marginal probability distribution $\gamma$ and hence the number of linked and unlinked cases isolated at time $t$ can be expressed as:

$$n_{comm,link,isolate}(t) = \int_0^{\infty} n_{comm,link}(t-\tau)\,\gamma(\tau)d\tau \qquad (4)$$
$$n_{comm,unlink,isolate}(t) = \int_0^{\infty} n_{comm,unlink}(t-\tau)\,\gamma(\tau)d\tau$$

We define the likelihood of observing unlinked and linked cases at time of isolation, $t_i$, as follows:

$$L_{t_i}^{offspring} = P_{pois}\left[observed_{\ comm,link,isolate}(t) \mid n_{comm,link,isolate}(t)\right] \times P_{pois}\left[observed_{\ comm,unlink,isolate}(t) \mid n_{comm,unlink,isolate}(t)\right] \qquad (5)$$

The final likelihood of the offspring infections over the course of the epidemic is as follows:

$$L = \prod_i L_{t_i}^{offspring} \qquad (6)$$

### Estimating Model Parameters

Given the long time series of data for model fitting, we subset the data, a priori, into five time periods when transmission was mainly driven by (i) Wuhan travellers, (ii) returning travellers from other countries with ongoing outbreak, followed by the implementation of (iii) lockdown within Singapore, (iv) resumption of local activities and (v) increased reopening of national borders.

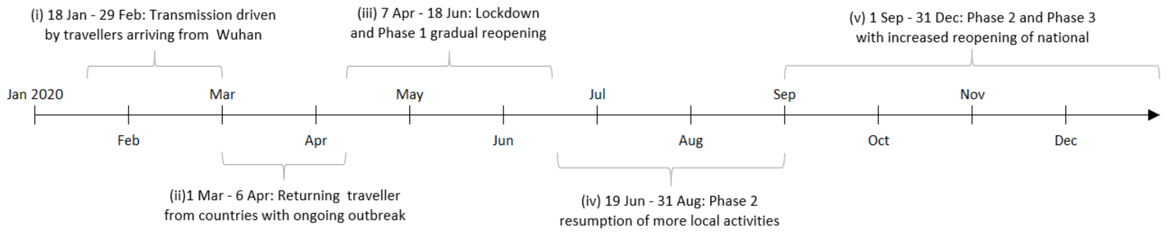


We assumed a uniform prior for the four model parameters $R, \varepsilon_{link}, \varepsilon_{unlink}$ and $\rho$ in each time period and multiple chains were run with a burn-in of 5,000 iterations and samples were thinned every 10 iterations. Convergence was assessed through visual inspection of the Gelman -Rubin convergence statistic and depending on the time period of interest, the posterior distribution of the parameters were estimated via Markov chain Monte Carlo sampling from 15,000 – 150,000 draws.

### Independent model validation

We estimate the number of number of missed infections infected between Apr 15 and May 14, 2020 that will be detected positive by PCR and serology from May 15 to 29, 2020 as follows:

$$\int_{t_{test\ start}}^{t_{test\ end}} \int_0^\infty \theta(t)m(t-\tau)\delta_{PCR}(\tau)S_+\delta_{Sero}(\tau)d\tau dt \tag{7}$$

where $\theta(t)$ is the empirical probability distribution of being tested on a day from May 15 to 29, 2020 (inclusive of both dates), $\delta_{PCR}$ is the probability of being detected positive by PCR $\tau$ time since infection, $S_+$ is the probability of seroconversion[3], $\delta_{Sero}$ is the probability of being detected serology positive $\tau$ time since infection given seroconversion.

$\delta_{PCR}$ and $\delta_{Sero}$ were derived by convolving the distribution of PCR and serology IgG detection probabilities since time of symptoms onset and the incubation period distribution for SAR-CoV-2 infections respectively.[1,4]

We crudely estimated the number of missed infections infected on or before Sep 30, 2020 that will be detected positive by serology IgG when tested by Oct 31, 2020 as follows:

$$\int_{30}^{\infty} m(t-\tau)S_+ d\tau \tag{8}$$

In other words, we assumed that $\delta_{Sero}(\tau)$ is equal to 1 when the time of infection was at least 30 days prior to serological testing given successful seroconversion.

Incidence rates per million population were derived by dividing the estimated number of missed infections positive on the respective test by 5.381 (i.e. the number of people in the general community was approximately 5 381 000).

**Relative role of $\varepsilon_{link}$, $\varepsilon_{unlink}$ and $\rho$ on the effective reproduction number, ratio of missed to notified cases, ratio of unlinked to linked cases and generations to exponential growth**

In a population with missed and notified SARS-CoV-2 parent infectors, the ability to identify the next generation of offspring infections are dependent on the efficacy of the contact tracing system in identifying secondary cases (i.e. probability of detecting linked cases, $\varepsilon_{link}$) and the efficacy of the surveillance system in case finding (i.e. probability of detecting unlinked cases, $\varepsilon_{unlink}$).

We define $k_{ij}$ to be the expected number of offspring infections of notification status $i$, caused by one parent infector with notification status $j$, during the time when the individual is at large in the community, and $i,j \in \{n,m\}$ where $n$ and $m$ denotes notified and missed respectively. There are four different transmission pairs and the next-generation matrix, $K$ can be expressed as:

$$K = \begin{pmatrix} k_{mm} & k_{mn} \\ k_{nm} & k_{nn} \end{pmatrix} \tag{9}$$

The computation of each element of $K$ is as follows:

- $k_{nm}$ denotes the expected number of notified offspring infections generated by a missed parent infector and can be expressed as $\varepsilon_{unlink}R$. These offspring infections are also termed as unlinked cases.

- $k_{mm}$ denotes the expected number of missed offspring infections generated by a missed parent infector and can be expressed as $(1-\varepsilon_{unlink})R$.

- $k_{nn}$ denotes the expected number of notified offspring infections generated by a notified parent infector and can be expressed as $\varepsilon_{link}R\int_0^i\int_0^\infty \gamma(i)\omega(\tau)\,did\tau$ . These offspring infections are also termed as linked cases. The reproduction number of a notified case is lower than a missed infection by a factor of $\int_0^i\int_0^\infty \gamma(i)\omega(\tau)\,did\tau$ as these cases are subjected to early isolation upon notification and prior to the end of their infectious period.

- $k_{nm}$ denotes the expected number of missed offspring infections generated by a notified parent infector and can be expressed as $(1-\varepsilon_{link})R\int_0^i\int_0^\infty \gamma(i)\omega(\tau)\,did\tau$ .

The next-generation of infectors are described as follows:

$$\phi_i^g = K\phi_j^{g-1} \tag{10}$$

where $\phi$ is a column vector of missed and notified infectors and the superscript denotes the generation of the infectors.

The dominant eigenvalue and eigenvector of matrix $K$ characterises the effective reproduction number and the ratio of missed to notified cases respectively. Figure 4 and 5a in the main text was generated by computing the dominant eigenvalue and eigenvector of varying next-generation matrices with user input values of $R$, $\varepsilon_{link}$ and $\varepsilon_{unlink}$. The ratio of the unlinked to linked cases shown in Figure 5b is derived by multiplying the elements of the eigenvector with $k_{nm}$ and $k_{nn}$.

To establish the epidemic growth dynamics initiated by varying ratios of missed and notified imported cases, we first define the transmission matrix for imported cases, $K_{import}$ as follows:

$$K_{import} = \begin{pmatrix} k_{mm_{import}} & k_{mn_{import}} \\ k_{nm_{import}} & k_{nn_{import}} \end{pmatrix} \tag{11}$$

where $m_{import}$ and $n_{import}$ denotes missed and notified imported cases. The computation of each element of $K_{import}$is as follows:

- $k_{nm_{import}}$ denotes the expected number of notified offspring infections generated by a missed imported parent infector and can be expressed as $\varepsilon_{unlink}R\int_a^\infty\int_0^\infty \alpha(a)\omega(\tau)\;dad\tau$. These offspring infections are also considered as unlinked cases. The reproduction number of a missed imported case is lower than a missed community infection by a factor of $\int_a^\infty\int_0^\infty \alpha(a)\omega(\tau)\;dad\tau$ as these cases spent part of their infectious period while overseas.

- $k_{mm_{import}}$ denotes the expected number of missed offspring infections generated by a missed imported parent infector and can be expressed as $(1-\varepsilon_{unlink})R\int_{a}^{\infty}\int_{0}^{\infty}\alpha(a)\omega(\tau)\ da d\tau$.

- $k_{nn_{import}}$ denotes the expected number of notified offspring infections generated by a notified imported parent infector and can be expressed as $\varepsilon_{link}R\int_{\tau=a}^{\tau=a+i}\int_{i=0}^{\infty}\int_{a=0}^{\infty}\alpha(a)\gamma_{import}(i)\omega(\tau)\, da di d\tau$ . These offspring infections are also considered as linked cases. The reproduction number of a notified imported case is lowered than a missed community infection by a factor of $\int_{\tau=a}^{\tau=a+i}\int_{i=0}^{\infty}\int_{a=0}^{\infty}\alpha(a)\gamma_{import}(i)\omega(\tau)\, da di d\tau$ as these cases spent part of their infectious period while overseas and are subjected to early isolation upon notification and prior to the end of their infectious period, and is expressed by the two inner integrals.

  $\gamma_{import}$ is the probability distribution function of the duration of arrival to isolation for imported cases with community exposure, obtained from the data.

- $k_{mn_{import}}$ denotes the expected number of notified offspring infections generated by a notified imported parent infector and can be expressed as $(1-\varepsilon_{link})R\int_{\tau=a}^{\tau=a+i}\int_{i=0}^{\infty}\int_{a=0}^{\infty}\alpha(a)\gamma_{import}(i)\omega(\tau)\, da di d\tau$.

The first and subsequent generations of community infectors are generated as follows:

$$\phi_i^1 = K_{import}\phi_{j_{import}} \tag{12}$$
$$\phi_i^g = K\phi_j^{g-1}, \qquad g \geq 2$$

**Supplementary Figures and Table**

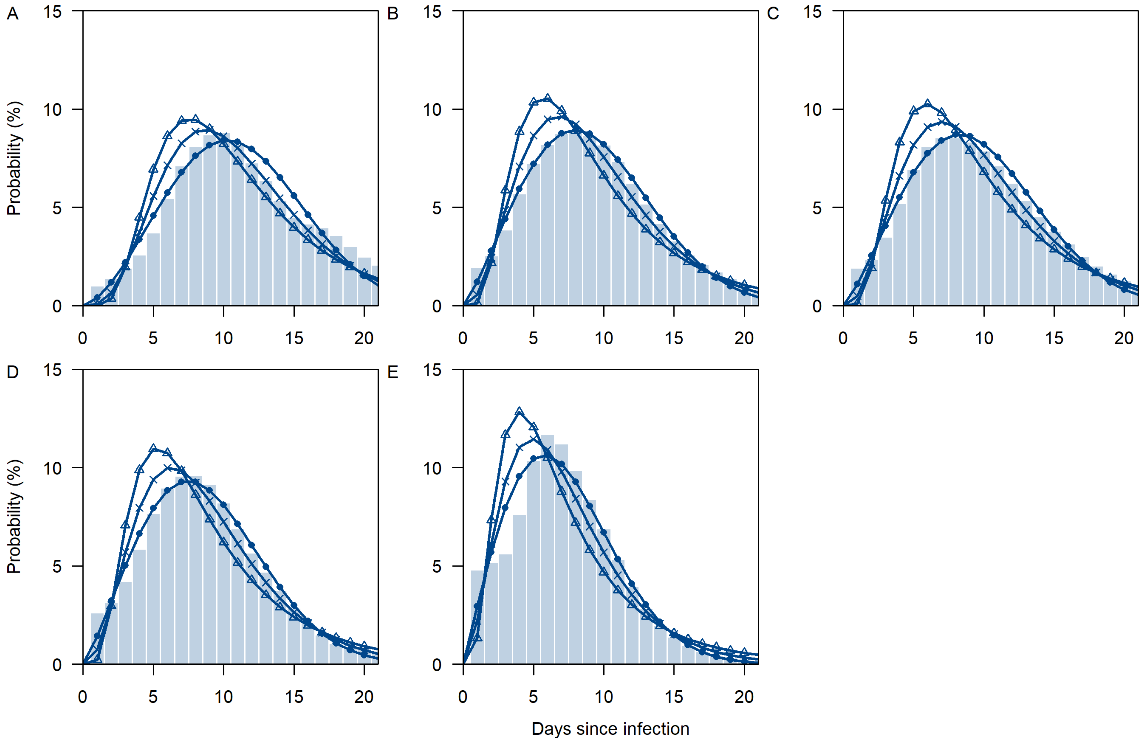

Supplementary Figure 1: Derived (bar) and model fit (dots: Weibull, cross: Gamma, triangle: Lognormal) of the marginal probability distribution of the duration of infection to isolation for community cases, $\gamma$. The derived distribution of $\gamma$ was obtained by convolving the observed distribution of time from symptoms onset in notified, symptomatic community cases to isolation, and the incubation period distribution for SAR-CoV-2 infections from (A) Jan 18 to Feb 29; (B) Mar 1 to Apr 6; (C) Apr 7 to Jun 18; (D) Jun 19 to Aug 31; (E) Sep 1, 2020 to Jan 1, 2021.

Supplementary Table 1: Model parameters for fitted $\gamma$ in respective time periods.

| Time period | Weibull (shape, scale) | Gamma (shape, rate) | Lognormal (meanlog, sdlog) |
|---|---|---|---|
| Jan 18 - Feb 29, 2020 | 2.56, 12.4 | 4.69, 0.42 | 2.29, 0.52 |
| Mar 1 - Apr 6, 2020 | 2.23, 10.4 | 3.70, 0.40 | 2.08, 0.59 |
| Apr 7 - Jun 18, 2020 | 2.24, 10.8 | 3.70, 0.39 | 2.11, 0.59 |
| Jun 19 - Aug 31, 2020 | 2.19, 9.85 | 3.59, 0.41 | 2.02, 0.60 |
| Sep 1, 2020 - Jan 1, 2021 | 2.01, 8.11 | 3.09, 0.43 | 1.80, 0.65 |

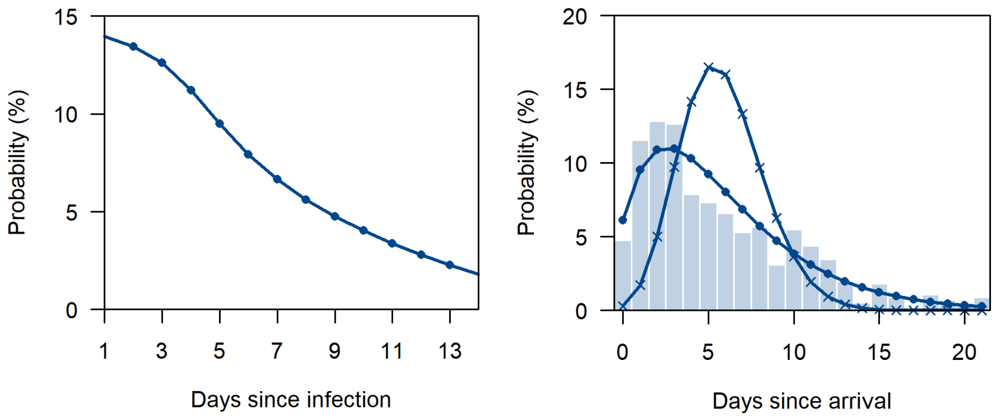


Supplementary Figure 2: Probability distribution of the duration of infection to arrival, $\alpha$ (left) and probability distribution of time of arrival to isolation in notified imported cases with community contact; (bar) observed data, (line) model fit (dots: negative binomial, cross: Poisson)

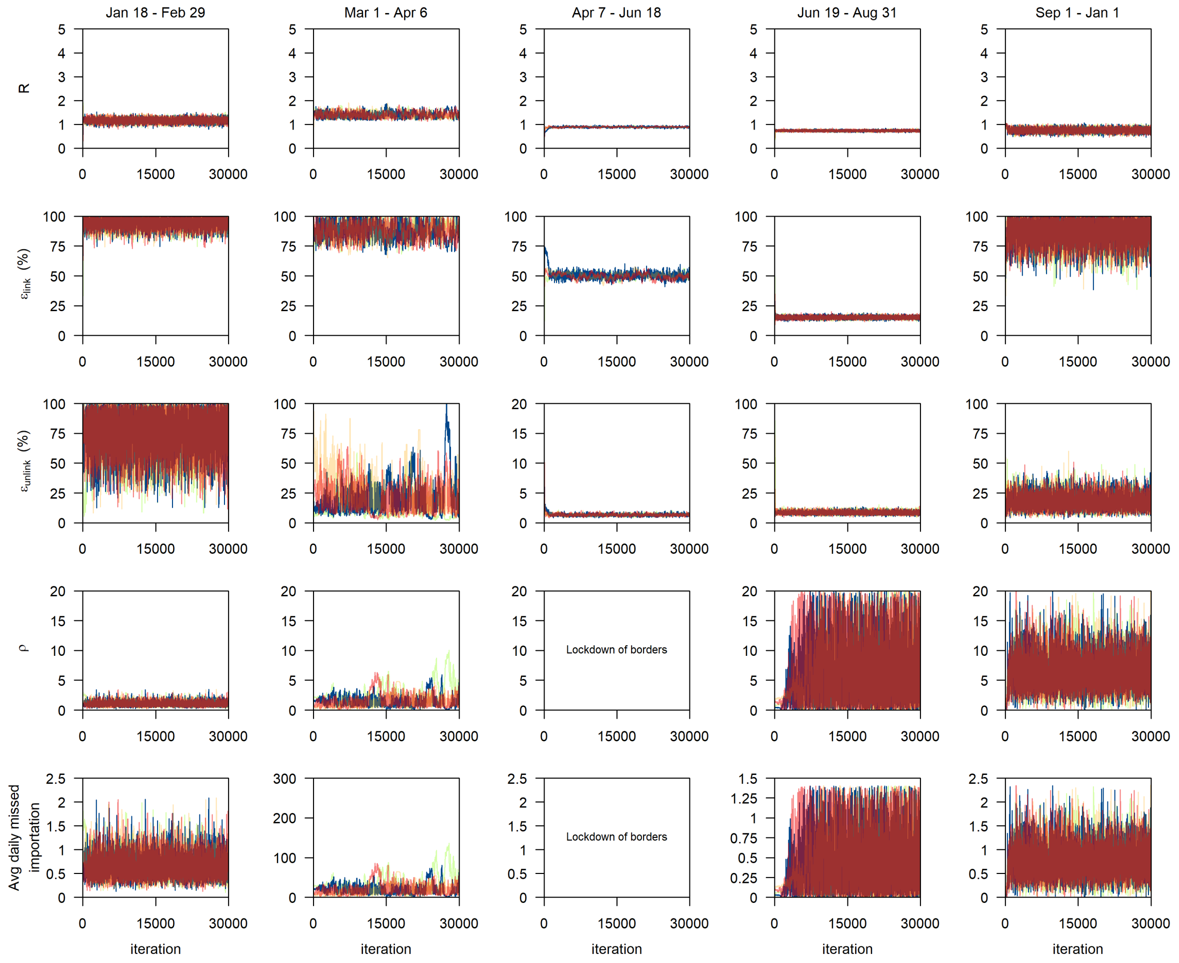


Supplementary Figure 3: Trace plot of model parameters, $R$ (first row), $\varepsilon_{link}$ (second row), $\varepsilon_{unlink}$ (third row) and $\rho$ (fourth row) for the first 30 000 iterations. The average daily missed importation (fifth row) is the derived from $\rho\bar{n}_{import}$ where $\bar{n}_{import}$ is the mean daily number of notified imported cases that arrived in the time period of interest.

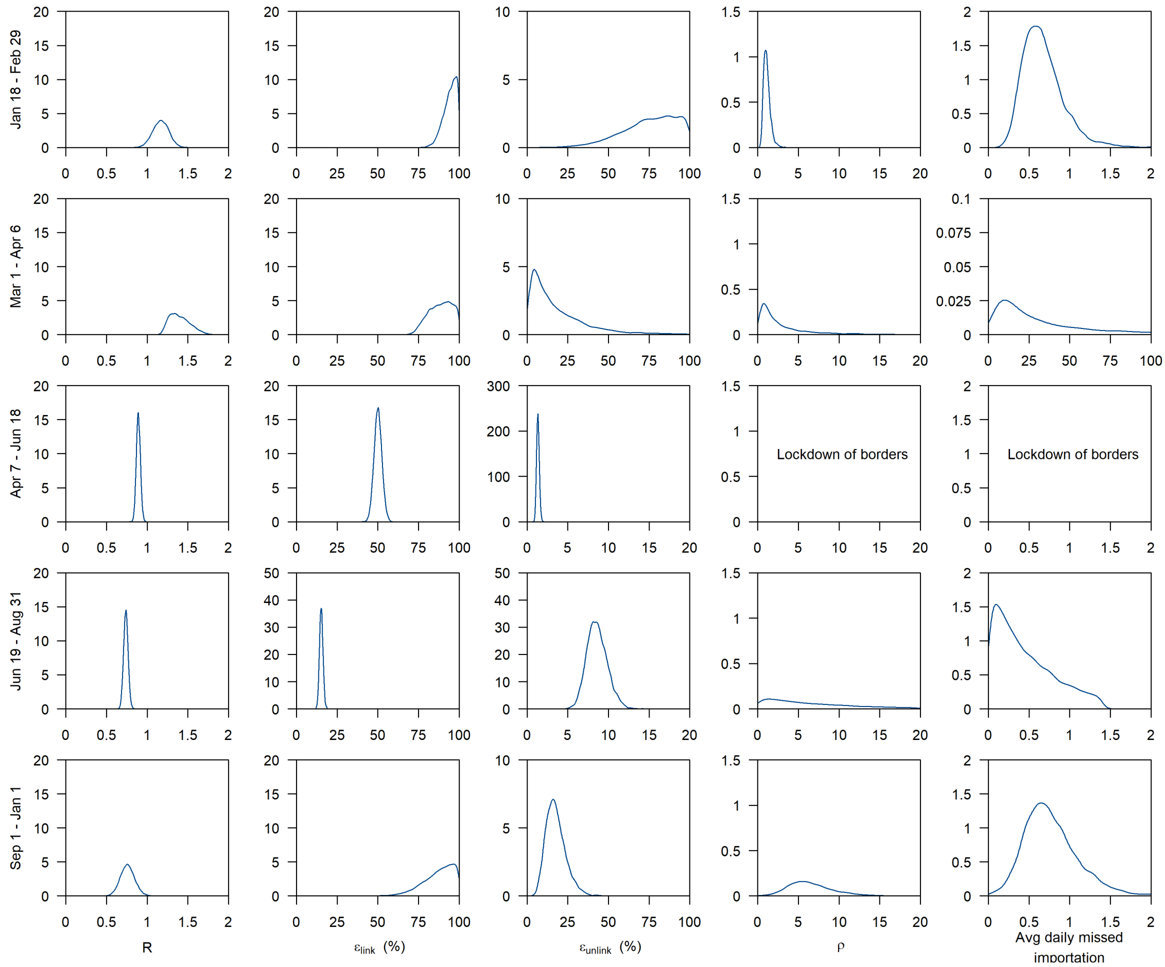


Supplementary Figure 4: Density plots of model parameters, $R$ (first column), $\varepsilon_{link}$ (second column), $\varepsilon_{unlink}$ (third column) and $\rho$ (fourth column). The average daily missed importation (fifth column) is the derived from $\rho\bar{n}_{import}$ where $\bar{n}_{import}$ is the mean daily number of notified imported cases that arrived in the time period of interest.

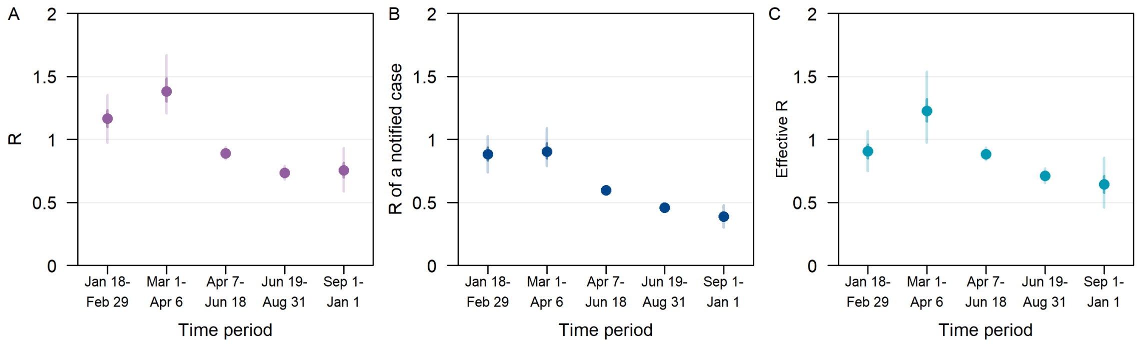


Supplementary Figure 5: Posterior median (dot), 50% CI (dark vertical lines) and 95% CI (light vertical lines) of (A) potential reproduction number, $R$, of a COVID-19 case for the entire infectious period in the absence of quarantine/isolation; (B) reproduction number of a notified

COVID-19 case, $R_{\text{notified}}$; (C) effective reproduction number, $R_{\text{eff}}$, after accounting for missed and notified COVID-19 infections.

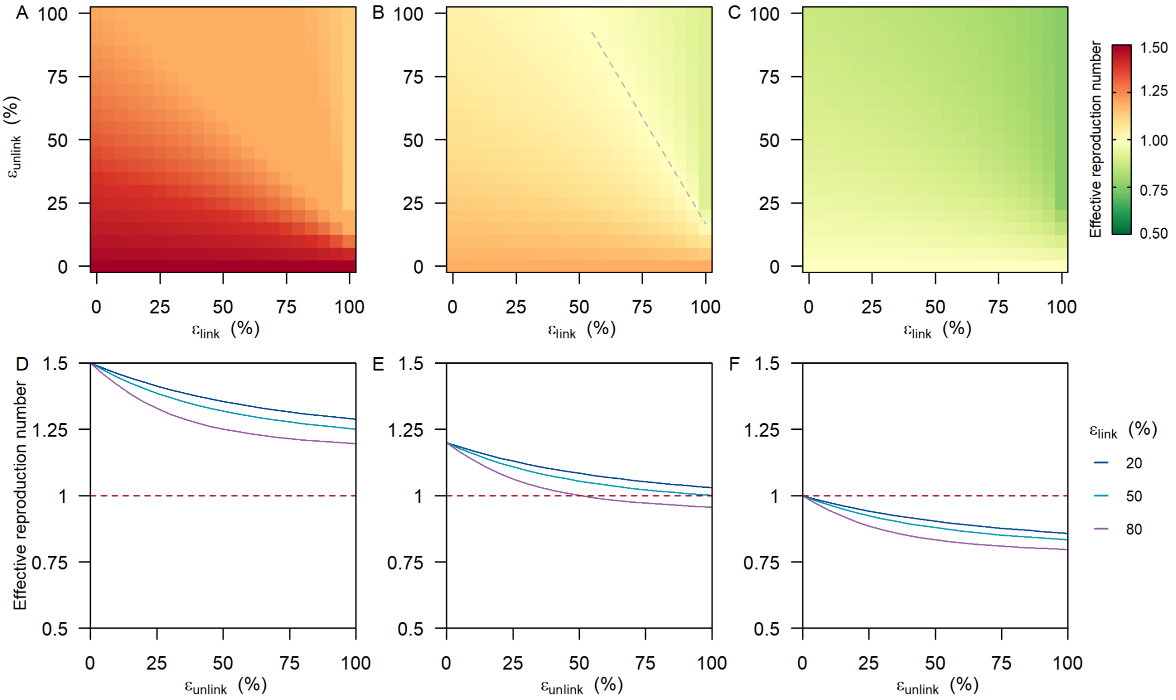


Supplementary Figure 6: Changes in effective reproduction number (i.e. $R_{\text{eff}}$) for a fixed distribution of time from infection to isolation and varying $\varepsilon_{\text{link}}$, $\varepsilon_{\text{unlink}}$ and potential reproduction number, $R$, of 1.5 (A, D), 1.2 (B, E) and 1.0 (C, F). The distribution of time from infection to isolation of notified cases follows a Weibull distribution with mean 11.0 days (SD 4.6), similar to the data observed between Jan 18 and Feb 29, 2021. (B) Grey dashed line represents effective R of 1.

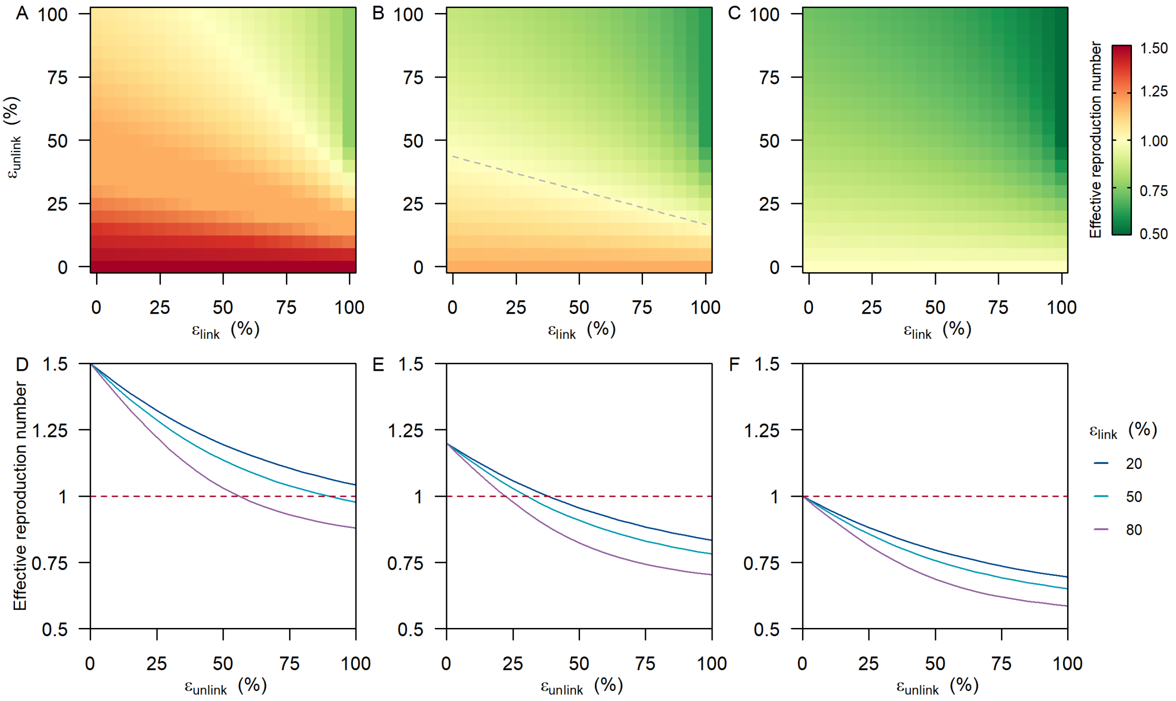


Supplementary Figure 7: Changes in effective reproduction number (i.e. $R_{\text{eff}}$) for a fixed distribution of time from infection to isolation and varying $\varepsilon_{\text{link}}$, $\varepsilon_{\text{unlink}}$ and potential reproduction number, $R$, of 1.5 (A, D), 1.2 (B, E) and 1.0 (C, F). The distribution of time from infection to isolation of notified cases follows a Weibull distribution with mean 7.2 days (SD 3.7), similar to the data observed between Sep 1, 2020 and Jan 1, 2021. (A, B) Grey dashed line represents effective R of 1.

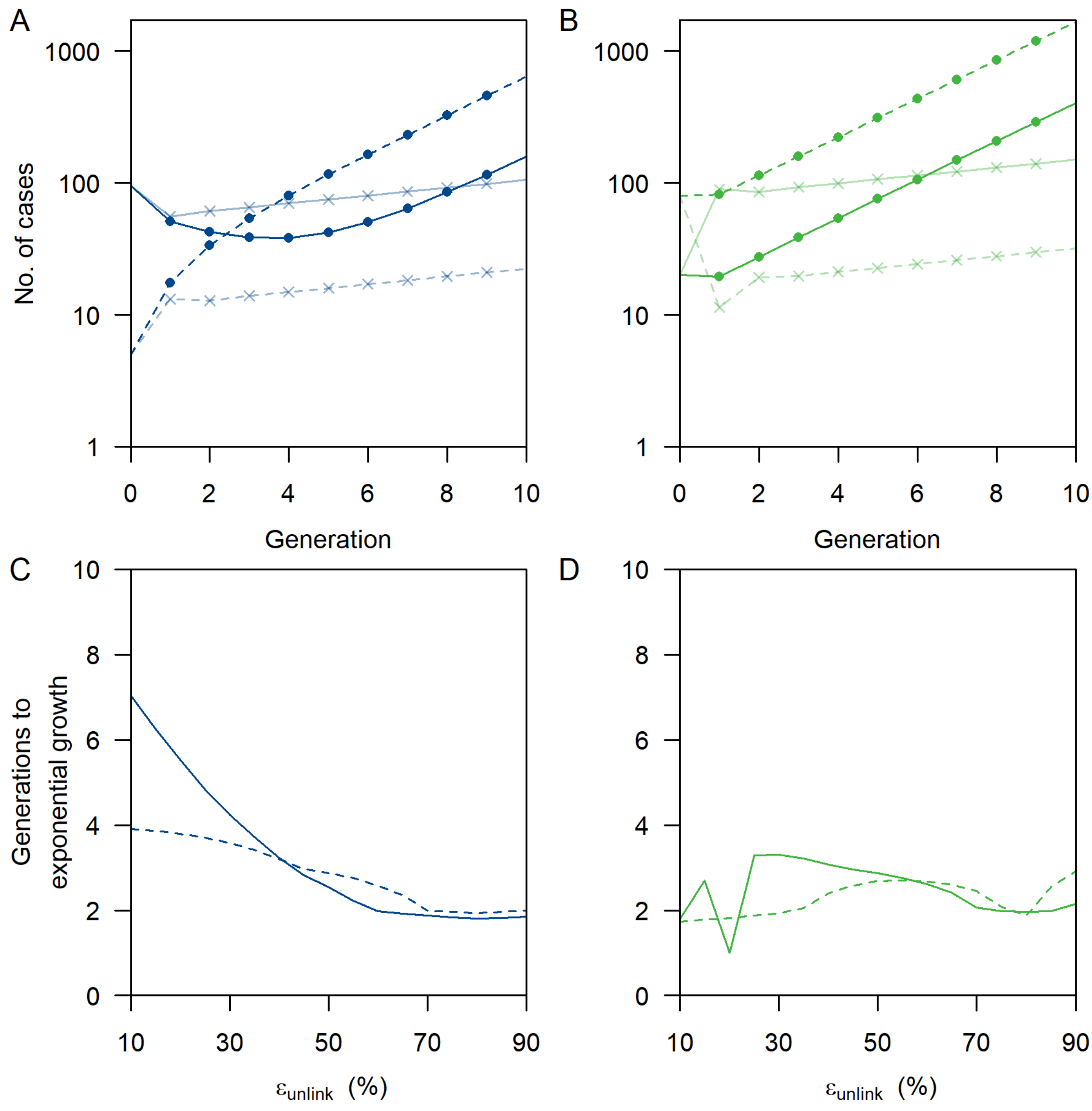


Supplementary Figure 8: Generations to exponential growth. Outbreak initiated with 5 missed imported cases (blue dashed line) and 95 notified imported cases with community contact (blue solid line), $R$ of 1.5, $\varepsilon_{link}$ of 80%, for (A) $\varepsilon_{unlink}$ of 10% (dots) and $\varepsilon_{unlink}$ of 90% (cross); (C) varying $\varepsilon_{unlink}$. Outbreak initiated with 80 missed imported cases (green dashed line) and 20 notified imported cases with community contact (green solid line), $R$ of 1.5, $\varepsilon_{link}$ of 80%, for (B) $\varepsilon_{unlink}$ of 10% (dots) and $\varepsilon_{unlink}$ of 90% (cross); (D) varying $\varepsilon_{unlink}$.


**The following authors were part of the Centre for Mathematical Modelling of Infectious Disease COVID-19 Working Group.** Each contributed in processing, cleaning and interpretation of data, interpreted findings, contributed to the manuscript, and approved the work for publication: Gwenan M Knight, David Hodgson, Graham Medley, Emily S Nightingale, Rosalind M Eggo, Matthew Quaife, C Julian Villabona-Arenas, Sebastian Funk, Christopher I Jarvis, Paul Mee, Billy J Quilty, Yalda Jafari, Yang Liu, Joel Hellewell, Hamish P Gibbs, Fabienne Krauer, James D Munday, Katherine E. Atkins, Damien C Tully, Ciara V McCarthy, Mihaly Koltai, Timothy W Russell, Kaja Abbas, Emilie Finch, W John Edmunds, Kiesha Prem, Nikos I Bosse, Sophie R Meakin, Frank G Sandmann, Yung-Wai Desmond Chan, Kevin van Zandvoort, Samuel Clifford, Kathleen O'Reilly, Naomi R Waterlow, Amy Gimma, Thibaut Jombart, Oliver Brady, Mark Jit, Simon R Procter, Rosanna C Barnard, Anna M Foss, William Waites, Fiona Yueqian Sun, Sam Abbott, Stefan Flasche, Nicholas G. Davies, Alicia Rosello, Akira Endo, Rachel Lowe, Carl A B Pearson, Jiayao Lei.


**The following funding sources are acknowledged as providing funding for the working group authors.** This research was partly funded by the Bill & Melinda Gates Foundation (INV-001754: MQ; INV-003174: JYL, KP, MJ, YL; INV-016832: SRP; NTD Modelling Consortium OPP1184344: CABP, GFM; OPP1139859: BJQ; OPP1183986: ESN; OPP1191821: KO'R). BMGF (INV-016832; OPP1157270: KA). CADDE MR/S0195/1 & FAPESP 18/14389-0 (PM). EDCTP2 (RIA2020EF-2983-CSIGN: HPG). Elrha R2HC/UK FCDO/Wellcome Trust/This research was partly funded by the National Institute for Health Research (NIHR) using UK aid from the UK Government to support global health research. The views expressed in this publication are those of the author(s) and not necessarily those of the NIHR or the UK Department of Health and Social Care (KvZ). ERC Starting Grant (#757699: MQ). ERC (SG 757688: CJVA, KEA). This project has received funding from the European Union's Horizon 2020 research and innovation programme - project EpiPose (101003688: AG, KP, MJ, RCB, WJE, YL). FCDO/Wellcome Trust (Epidemic Preparedness Coronavirus research programme 221303/Z/20/Z: CABP, KvZ). This research was partly funded by the Global Challenges Research Fund (GCRF) project 'RECAP' managed through RCUK and ESRC (ES/P010873/1: CIJ, TJ). HDR UK (MR/S003975/1: RME). HPRU

(NIHR200908: NIB). Innovation Fund (01VSF18015: FK). MRC (MR/N013638/1: EF, NRW; MR/V027956/1: WW). Nakajima Foundation (AE). NIHR (16/136/46: BJQ; 16/137/109: BJQ, FYS, MJ, YL; 1R01AI141534-01A1: DH; Health Protection Research Unit for Modelling Methodology HPRU-2012-10096: TJ; NIHR200908: AJK, RME; NIHR200929: CVM, FGS, MJ, NGD; PR-OD-1017-20002: AR, WJE). Royal Society (Dorothy Hodgkin Fellowship: RL). Singapore Ministry of Health (RP). UK DHSC/UK Aid/NIHR (PR-OD-1017-20001: HPG). UK MRC (MC_PC_19065 - Covid 19: Understanding the dynamics and drivers of the COVID-19 epidemic using real-time outbreak analytics: NGD, RME, SC, TJ, WJE, YL; MR/P014658/1: GMK). Authors of this research receive funding from UK Public Health Rapid Support Team funded by the United Kingdom Department of Health and Social Care (TJ). UKRI Research England (NGD). UKRI (MR/V028456/1: YJ). Wellcome Trust (206250/Z/17/Z: AJK, TWR; 206471/Z/17/Z: OJB; 208812/Z/17/Z: SC, SFlasche; 210758/Z/18/Z: JDM, JH, SA, SFunk, SRM; 221303/Z/20/Z: MK; UNS110424: FK). No funding (AMF, DCT, YWDC).
